# A systematic review and analysis of health risks associated with zootherapeutic practices in Africa

**DOI:** 10.1101/2024.02.14.24302750

**Authors:** Léa Fourchault, Abdallah Lamane, Dimitri Romaric Nguinwa Mbakop, Ganiyat Temidayo Saliu, Sophie Gryseels, Erik Verheyen, Katharina Kreppel

## Abstract

**Background:** Over five billion people globally rely on a plant- and animal-based pharmacopoeia for their healthcare needs. The inhalation, topical application, subcutaneous injection, or ingestion of animal products – such as faeces, fur, milk, blood, brain tissue, or meat – likely facilitates the spill over of zoonotic pathogens. Certain practices use species known to be involved in the transmission of pathogens of public health relevance, such as reservoir species for filoviruses, poxviruses, and coronaviruses. These practices and the public health risk they entail have not been previously reviewed and analysed for the African continent.

**Methods:** We first conducted a systematic review of literature using web-scraping algorithms targeted at peer-reviewed (PubMed) and peer-reviewed or grey literature (Google Scholar) databases, followed by manual search of reference lists published before July 30th, 2023. We used terms encompassing zoo*, animal*, health*, practice*, tradition* followed by a list of all 54 African countries in combination with Boolean operators. We then created a categorical score reflecting the risk of zoonotic pathogen spill over for each recorded zootherapeutic practice, and compared this risk between geographic regions and between demographic groups.

**Findings:** A total of 53 studies were included, reporting the use of over 2,000 zootherapeutic practices. Half of the included studies were published after 2020. Nigerian, Ethiopian, Tanzanian and South African practices were comparatively well documented. The mean total risk score was significantly lower in western (13.27 ± 0.13, p < 0.0001), central (14.80 ± 0.27, p < 0.003), and southern (13.48 ± 0.23, p < 0.0001) Africa, compared to eastern Africa, while there was no significant difference between eastern and northern Africa (15.25 ± 0.26, p = 0.16). Further, we found that physically sick children are overall at increased risk for pathogen spill over (13.20 ± 0.36 out of a possible sub-score of 20, p = 0.001 < 0.05) compared to physically sick adults, and that pregnant or lactating women are exposed to animal tissues of significantly greater infectious potential (4.01 ± 0.15, p = 0.032 <0.05).

**Interpretation:** The WHO recently hosted its first global summit on Traditional, Complementary and Integrative Medicine (TCIM), highlighting its importance to fulfil SDG 3: Good Health and Wellbeing. Where other forms of healthcare are unavailable or inefficient, zootherapeutic practices can provide valuable solutions to acute, chronic, physical, and psychological issues. However, significant risks of zoonotic disease transmission exist. This article aims to guide research on sustainable alternatives to mainstream medical treatments that balance cultural significance and public health.

## Introduction

Both wild and domestic animals host pathogens that can spill over to humans (Karesh *et al*., 2012; Carlson *et al*., 2021; Grange *et al*., 2021; Gamble *et al*., 2023). While most spill over events are inconsequential, some have major impacts on public health and economies (Cutlers and Summers, 2020). Examples include several HIV strains that originally spilled over from African primates (Peeters *et al*., 2014), Mpox spilling over from African mammals (with the specific reservoir(s) still being investigated, e.g., Falendysz *et al*., 2023), or *Brucella* bacteria from cattle, a predominant issue in northern and eastern Africa (Mburu *et al*., 2020; Djaafri *et al*., 2022). So far, hunting, and the butchering and consumption of wild and domestic animal meat have been investigated as major mechanisms for spill over, while other transmission routes, such as zootherapy, remain under-researched (Paige *et al*., 2014; Friant *et al*., 2022; Milbank and Vira, 2022; Moloney *et al*., 2023).

Five billion people primarily rely on Traditional, Complementary, and Integrative Medicine (TCIM) for their healthcare and wellbeing, including about 80% of the over one billion people inhabiting Africa (Albertyn *et al*., 2015). Zootherapy, the use of animal *materia medica* (e.g., fur, excreta, bones, blood), is an integral part of TCIM (Alves *et al*., 2011). Zootherapeutic practices are a major source of exposure to animal products and the pathogens they may carry (Alves *et al*., 2021; Friant *et al*., 2022). Each topical application, injection, inhalation, or ingestion of such animal products is therefore a potential mechanism for spill over (Plowright *et al*., 2017; Friant *et al*., 2022). Identifying zootherapeutic practices of higher epidemiological risk is thus crucial to develop sustainable alternatives that balance cultural significance and public health.

Our first aim was to analyse geographic and temporal trends in the recording of African zootherapeutic practices since 1990, to guide future research. Our second aim was to characterise geographic and demographic variations in the risk of zoonotic pathogen spill over, based on animal tissue types used as *materia medica*, methods of treatment administration, level of gregariousness of the animal, phylogenetic relatedness between the animal used and human patients, and immunocompetency levels of the patients exposed to zootherapeutic practices.

## Methods

### Meta-analysis

Following the updated PRISMA guidelines (Page *et al*., 2020), we conducted a systematic review of literature published until 30^th^ of July 2023, using web-scraping algorithms targeted at peer-reviewed (PubMed) and peer-reviewed or grey literature (Google Scholar) databases, followed by manual search of reference lists. We also obtained publications and master/doctoral theses from the main organisation affiliated with this study (Royal Belgian Institute of Natural Sciences). Terms encompassing zoo*, animal*, health*, practice*, tradition* followed by a list of all 54 African countries were used in combination with Boolean operators. Studies were included if they were published in peer-reviewed articles or as scientific theses in French or English after 1990, explicitly stated the name of the animal and the ailment treated, the animal tissue type used, and/or the treatment method. Three independent reviewers retrieved (AL, LF, GTS), screened (LF, GTS) and assessed (LF, GTS) the studies. In addition to the extraction of metadata, a data extraction template was created with five main sections, focusing on: the animal (species, genus, order, class, phylogenetic relatedness, level of gregariousness), the tissue type, the method of treatment administration, and the human patient. Our study was not registered with PROSPERO.

We then investigated the geographic and temporal variations in the recorded zootherapeutic practices by mapping the number of studies and total study size in each country, and by constructing saturation curves of the cumulative number of distinct practices recorded with each new study.

### Selection of the risk factors and risk analysis

We first assessed the risk of zoonotic pathogen spill over for each distinct zootherapeutic practice using a scoring system that reflects the likelihood of the successful establishment of any pathogen in the human patient after performing this zootherapeutic practice. Using peerreviewed evidence, we first identified risk factors suggested to contribute to zoonotic spill over risk, which is characterized by the ability of an animal-sourced virus to infect and cause disease in humans (Grange *et al*., 2021).

Commonly cited risk factors included the phylogenetic relatedness between the animal species and humans (Dharmarajan *et al*., 2022; see Becker *et al*., 2019 for nuances), the level of gregariousness of the animal species (Dharmarajan *et al*., 2022), and the immunocompetence of the human recipient at the time of the treatment (Simon *et al*., 2015; Plowright *et al*., 2017). We also chose to include the animal tissue type that the patient would be in contact with (e.g., bones, fur, blood), and the method of treatment administration (e.g., ingestion after cooking, inhalation, topical application on a wound), as pathogens are present at higher concentrations in certain tissues, and are more likely to reach human cells through certain exposure routes (Plowright *et al*., 2017). We then scored the components making up each of these risk factors, from five (highest risk of zoonotic pathogen spill over) to one (lowest risk), as shown in Table 1. The total risk score was created by summing the scores across factors for each practice. To examine the impact of each factor, we used a sensitivity analysis where factors were given different weights.

**Table 1.** Categorical components of the risk factors. Unless otherwise specified, all components were weighted equally. ‘Altered’ can be dried, sun-dried, crushed to powder, smoked, or a combination of these. ‘Spraying/pouring’ refers to animal tissues being sprayed or poured around the patient (e.g., on the walls or floor).

| **Factor/**<br>**Score** | **Tissue type** | **Treatment type** | **Phylogenetic**<br>**relatedness** | **Gregariousness** | **Immunocompetence**<br>**(Human recipient)** |
| --- | --- | --- | --- | --- | --- |
| **Highest**<br>**risk (5)** | Blood,<br>internal organs,<br>whole animal<br>(including<br>foetus) | Injected, topical<br>(raw) on wound | Primates | Social | Physically sick child<br>or infant |
| **High**<br>**risk (4)** | Faeces, urine,<br>secretions<br>(vaginal,<br>seminal,<br>saliva), milk,<br>flesh, fat, skin,<br>eggs | Spraying/pouring<br>inhalation (raw),<br>ingestion (raw),<br>topical (raw) on<br>mucosa, topical<br>(altered/cooked)<br>on wound | Other<br>mammals | NA | Seemingly physically<br>healthy child or infant<br>(psychological or<br>spiritual issues, or<br>preventive medicine) |
| **Medium**<br>**risk (3)** | Bones | Ingestion<br>(altered), topical,<br>topical<br>(altered/cooked)<br>on mucosa | Birds | Restricted family<br>unit or<br>seasonally social | Pregnant or lactating<br>woman |
| **Low risk**<br>**(2)** | Hair/fur,<br>feathers | Inhalation<br>(altered) | Reptiles,<br>amphibians,<br>fish | NA | Physically sick adult |
| **Lowest**<br>**risk (1)** | Honey, butter,<br>scales, nails,<br>horns, sting,<br>venom, teeth | Ingestion<br>(cooked) | Invertebrates | Solitary | Seemingly physically<br>healthy adult<br>(psychological or<br>spiritual issues, or<br>preventive medicine) |

We then used Generalized Linear Models to analyse the distribution of this risk score. To examine the risk score across geographic regions, we used the total risk score, where all five factors are included (maximal possible total score of 25). To examine the risk score across demographic groups, we used a sub-score that excludes immunocompetency, as immunocompetency correlates with the demographic categories of interest for this study (maximal possible sub-score of 20). Finally, we examined the correlation between each risk factor and each demographic category, to characterize the driving factors of risk for vulnerable demographics (*i*.*e*., children/infants, and pregnant or lactating women).

## Results

Of the 2031 records retrieved, 415 full-texts were assessed for eligibility and 53 studies met all the inclusion criteria (Figure 1, Table S1).

**Figure 1.**
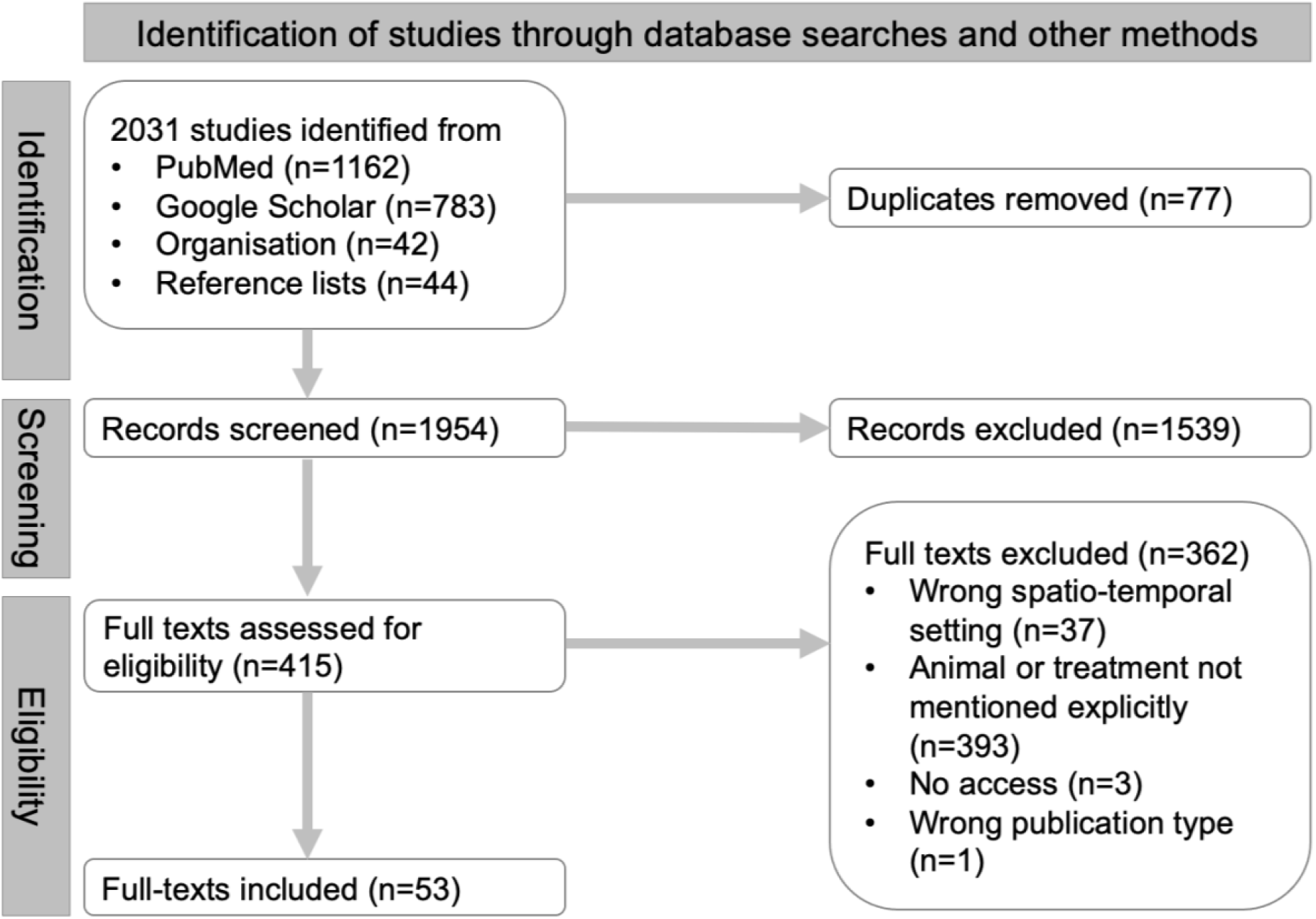
PRISMA flow chart highlighting the search outcomes. Following the updated guidelines for study selection (Page *et al*., 2020).

Half of the 53 included studies were published in or after 2020, leading to a stark temporal increase in the number of newly recorded practices, and 37 studies were led by African scholars (Figure 2A, Table S1). With ten studies published in the last thirty years, Nigeria had both the highest number of publications and the highest overall study size (Figure 2B, 2C and 2D). The saturation curve indicates that the number of new practices recorded in each new study started to plateau in Nigeria, with no plateau in other countries yet (Figure 2B).

**Figure 2.**
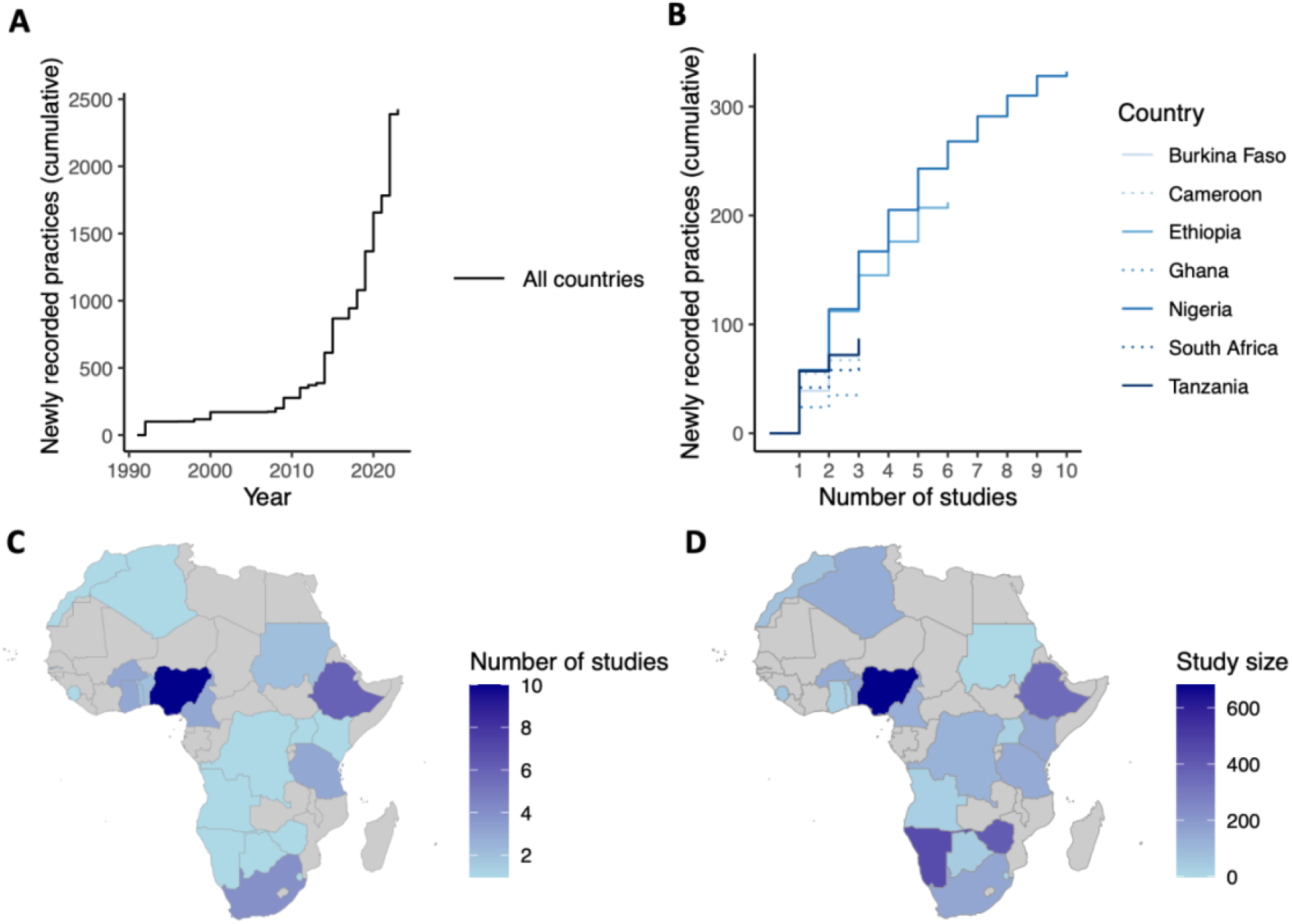
Spatial and temporal trends in studies on African zootherapeutic practices. A: number of new practices recorded since 1990. B: saturation curve representing the number of new practices recorded with each new study (the graph shows a subset of the seven countries having the greatest number of studies). C: number of studies published for each country. D: overall study size for each country (all studies included).

The mean total risk of zoonotic pathogen spill over was moderate (14.00 ± 2.80 out of a maximum possible total score of 25, n = 2425, Figure S2), regardless of the weight given to factors of potentially lower importance (Figures S3) or the focal ailment (Figure S4). We found significant regional differences in the risk of zoonotic pathogen spill over. The mean total risk score was significantly lower in western (13.27 ± 0.13, p < 2e-16, n = 1456), central (14.80 ± 0.27, p = 0.00279 < 0.05, n = 108), and southern (13.48 ± 0.23, p < 2e-16, n = 165) Africa, compared to eastern Africa (15.62 ± 0.11, n = 563), while there was no significant difference between eastern and northern Africa (15.25 ± 0.26, p = 0.16 > 0.05, n = 118; Figure 3A). Phylogenetic relatedness between humans and animals used as the source of the treatment, as well as the method of treatment, were major contributors to the total risk in most countries. Exceptions include Benin, where phylogenetic relatedness minimally contributed to risk; and D.R. Congo and Zimbabwe, where the method of treatment minimally contributed to risk (Figure 3B).

**Figure 3.**
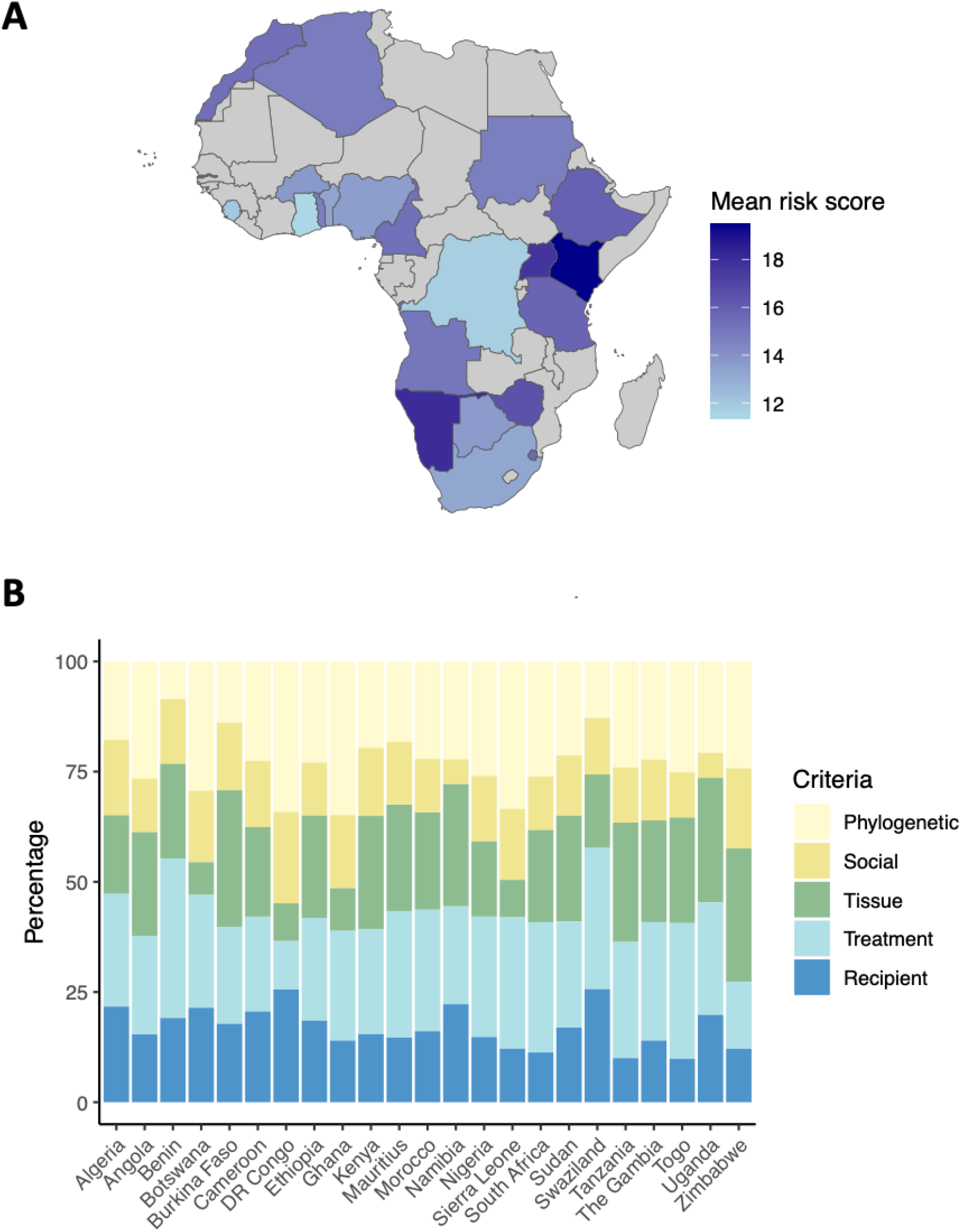
Spatial distribution of risk. A: risk map. B: stacked bar plot showing the drivers of risk (%) for each country. DR Congo: Democratic Republic of Congo.

Last, we found that the mean risk of zoonotic pathogen spill over across all demographics, as indicated by the sub-score, was significantly higher for physically sick children than for other patient categories (13.20 ± 0.36 out of a possible sub-score of 20, p = 0.001 < 0.05, Table 2), but not for pregnant or lactating women (12.09 ± 0.31, p = 0.83 > 0.05). The detailed impact of each factor on the risk of pathogen spill over by patient category of interest highlights a significantly higher risk for physically sick children and for children subjected to zootherapeutic practices for spiritual, psychological, or preventive purposes, with two out of four factors carrying a higher risk (e.g., riskier treatment methods: 2.67 ± 0.14, p = 0.000785 <0.05). While not overall more at risk, pregnant or lactating women are exposed to animal tissues of significantly greater infectious potential (4.01 ± 0.15, p = 0.032 <0.05). Finally, adults subjected to zootherapeutic practices for spiritual, psychological, or preventive purposes are exposed to animals of greater phylogenetic relatedness, such as primates or other mammals (3.61 ± 0.06, p = 3.84^-10^ > 0.05).

**Table 2.** Risk and risk drivers across demographic categories. The p-value is given in brackets after each estimate, in bold if the p-value is significant or approaching significance, and the estimate is positive (higher risk). Reference group: physically sick adults. The subscore (‘risk score’) used here excludes immunocompetence, which correlates with the human patient category. ‘Seemingly physically healthy’ refers to psychological, spiritual, or preventive zootherapeutic practices, as described in Table 1.

| Factor/<br>Human<br>patient | Risk score<br>(p-value) | Tissue type | Treatment<br>type | Phylogenetic<br>relatedness | Gregariousness |
| --- | --- | --- | --- | --- | --- |
| **Physically**<br>**sick adult** | 12.02 | $3.74 \pm 0.03$ | $2.19 \pm 0.02$ | $3.23 \pm 0.03$ | $2.86 \pm 0.04$ |
| **Seemingly**<br>**physically**<br>**healthy**<br>**adult** | $-0.05 \pm 0.14$<br>(0.69460) | $+0.03 \pm 0.07$<br>(0.637) | $-0.26 \pm 0.05$<br>($1.08e-06$ ***) | $+0.38 \pm 0.06$<br>**($3.84e-10$ \*\*\*)** | $-0.20 \pm 0.09$<br>(0.0330 *) |
| **Pregnant**<br>**or**<br>**lactating**<br>**woman** | $+0.07 \pm 0.31$<br>(0.82774) | $+0.32 \pm 0.15$<br>**($0.032$ \*)** | $-0.28 \pm 0.12$<br>(0.023545 *) | $+0.11 \pm 0.14$<br>(0.412723) | $-0.08 \pm 0.21$<br>(0.6927) |
| **Seemingly**<br>**physically**<br>**healthy**<br>**child** | $+0.63 \pm 0.38$<br>**($0.09511$ .)** | $-0.28 \pm 0.18$<br>(0.125) | $-0.05 \pm 0.15$<br>(0.761115) | $+0.47 \pm 0.17$<br>**($0.004910$ \*\*)** | $+0.49 \pm 0.26$<br>**($0.0603$ .)** |
| **Physically**<br>**sick child** | $+1.18 \pm 0.36$<br>**($0.00106$ \*\*)** | $-0.04 \pm 0.17$<br>(0.813) | $+0.48 \pm 0.14$<br>**($0.000785$ \*\*\*)** | $+0.54 \pm 0.16$<br>**($0.000673$ \*\*\*)** | $+0.21 \pm 0.25$<br>(0.3989) |

## Discussion

Our analysis first aimed at identifying trends in the publication landscape regarding primary research on zootherapeutic practices. We found that this topic is gaining traction, with half of the included studies being published in the last three years. Likely drivers of this surge in interest encompass recently increased research efforts in the domain of zoonoses, aimed at a better understanding of spill over mechanisms (e.g., Friant *et al*., 2022); and the increased attention given to TCIM to reach the Sustainable Development Goal 3: Good Health and Wellbeing (Patwardhan *et al*., 2023). Except for Nigeria, no country has yet reached a plateau in terms of new practices reported, indicating the need for more research. While research should be encouraged in all African countries, the Sahel region, parts of central Africa, and parts of east Africa entirely lack recent, peer-reviewed data on currently used zootherapeutic practices and should thus be considered priority regions for such studies.

Second, our analysis aimed at uncovering demographic and geographic trends in pathogen spill over risk associated with zootherapeutic practices. Our results indicate that vulnerable population members, especially children, are at an overall greater risk of zoonotic pathogen spill over through zootherapeutic practices than adults, mostly because of treatment methods of greater infectious potential. Examples include the injection of fluids containing *Mandrillus* sp. bones or *Gorilla* sp. bones to prevent “weakness” in children. Further, pregnant or lactating women are significantly more exposed to animal tissues of greater infectious potential, as, for instance, through the topical application of *Oryctolagus* sp. blood to treat chloasma, or the ingestion of *Rattus* sp. faeces to avoid childbirth complications. Given variations in the immunocompetency of these demographic groups (Simon *et al*., 2015), paediatric and maternal care practices should be carefully monitored.

Further, the inferred risk score was significantly lower in western, central, and southern Africa, compared to eastern Africa. This observation may be linked to the frequent use of animal products from domesticated animals in eastern Africa, which provide easy access to tissues of greater infectious potential, such as milk, blood, semen, or saliva. Veterinary services in eastern Africa therefore have an important role to play, among others in controlling brucellosis (Mburu *et al*., 2020). Similar comments can be made for north-African countries (Djaafri *et al*., 2022).

Last, we wish to highlight that the risks linked to zootherapeutic practices are exacerbated by at least two other factors: antimicrobial resistance, and hazardous preservatives. Multiple strains of antimicrobial-resistant bacteria have already been found in animal *materia medica*, for instance *Pseudomonas* sp. and *Salmonella* sp. in cow urine used for zootherapy in Nigeria (Ogunshe *et al*., 2010). Further, hazardous chemicals used to preserve animal *materia medica* may also have deleterious impacts on health (Zanvo *et al*., 2021). It is thus imperative to assess the risks and benefits of TCIM in a more holistic manner for TCIM to genuinely contribute to better health globally.

## Data availability

All data will be made available by the lead author upon request, and the code will be deposited into a publicly available repository on Github after publication.

## Notes

### Competing Interest Statement

The authors have declared no competing interest.

### Funding Statement

This study did not receive any funding.

