## Supplementary material for "A systematic review and analysis of health risks associated with zootherapeutic practices in Africa": Table S1: Supplemental information final.pdf

**Table S1. Included studies and their characteristics.**

| Country | Date | Authors | Local affiliation (lead author or PI) |
|---|---|---|---|
| Algeria | 2012 | Volpato et al. | None |
| Angola | 2017 | Braga-Pereira et al. | None |
| Benin | 2018 | Dossou et al. | Both |
| Benin | 2019 | Loko et al. | Both |
| Botswana | 2014 | Setlalekgomo | Both |
| Burkina Faso | 2007 | Beiersmann et al. | None |
| Burkina Faso | 2020 | Ouoba et al. | Both |
| Burkina Faso | 2022 | Ouango et al. | Both |
| Cameroon | 1996 | De Rosny | Both |
| Cameroon | 2015 | Emmanuel & Mamoudou | Both |
| Cameroon | 2015 | Bobo et al. | Both |
| DR Congo | 2020 | Mapoli Mbusa et al. | Both |
| Ethiopia | 2014 | Carruth | None |
| Ethiopia | 2020 | Tesfaye & Erena | Both |
| Ethiopia | 2022 | Kumera et al. | Both |
| Ethiopia | 2022 | Biru et al. | Both |
| Ethiopia | 2022 | Wendimu & Tekalign | Both |
| Ethiopia | 2023 | Birhan | Both |
| Ghana | 2011 | Insoll | None |
| Ghana | 2015 | Boakye et al. | None |
| Ghana | 2021 | Steinhorstel et al. | None |
| Kenya | 2017 | Riang'a et al. | Both |
| Mascarene Archipelago | 2013 | Mootoosamy & Mahomoodally | Both |
| Mascarene Archipelago | 2019 | Mahomoodally et al. | Both |
| Morroco | 2021 | Budjaj et al. | None |
| Namibia | 2022 | Uushona et al. | Both |
| Nigeria | 1992 | Adeola et al. | Both |
| Nigeria | 2008 | Soewu | Both |
| Nigeria | 2009 | Soewu and Ayodele | Both |
| Nigeria | 2009 | Eze et al. | Both |
| Nigeria | 2011 | Soewu and Adekanola | Both |
| Nigeria | 2017 | Ajagun et al. | Both |
| Nigeria | 2018 | Alade et al. | Both |
| Nigeria | 2020 | Gurumyen et al. | Both |
| Nigeria | 2022 | Zainab Muhammad et al. | Both |
| Nigeria | 2022 | Friant et al. | None |
| Sierra Leone | 2014 | Boakye | None |
| South Africa | 2016 | Williams & Whiting | Both |
| South Africa | 2019 | Nieman et al. | Both |
| South Africa | 2019 | Zondi | Both |
| South Africa | 2021 | Mashele et al. | Both |
| Subsaharan Africa | 2021 | Svensson et al. | None |
| Sudan | 2000 | El-Khamali | Both |

| | | | |
|---|---|---|---|
| Sudan | 2020 | Sumia et al. | Both |
| Swaziland (eSwatini) | 2021 | Nann | None |
| Tanzania | 2015 | Vats & Thomas | Both |
| Tanzania | 2015 | Magige | Both |
| Tanzania | 2018 | Roulette et al. | PI only |
| The Gambia | 1998 | Madge | None |
| Togo | 2020 | D'Cruze et al. | None |
| Togo | 2021 | Assou et al. | Lead author only |
| Uganda | 2021 | Isabirye | Both |
| Zimbabwe | 2019 | Mawoza et al. | Both |

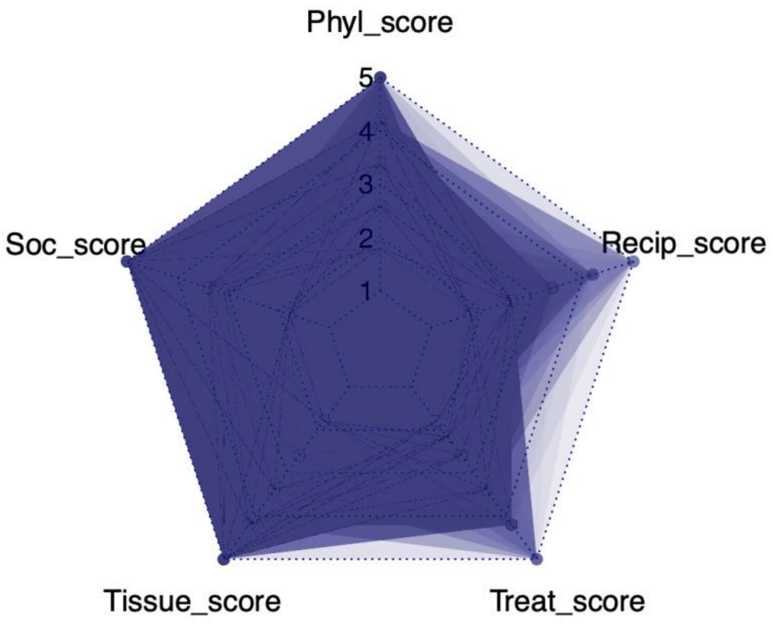


**Figure S2. Visual distribution of the risk score components.** 'Phyl': phylogenetic relatedness, 'Recipt': immunocompetence of the patient, 'Treat': treatment administration method, 'Tissue': tisssue type of the animal, 'Soc': level of gregariousness of the animal.

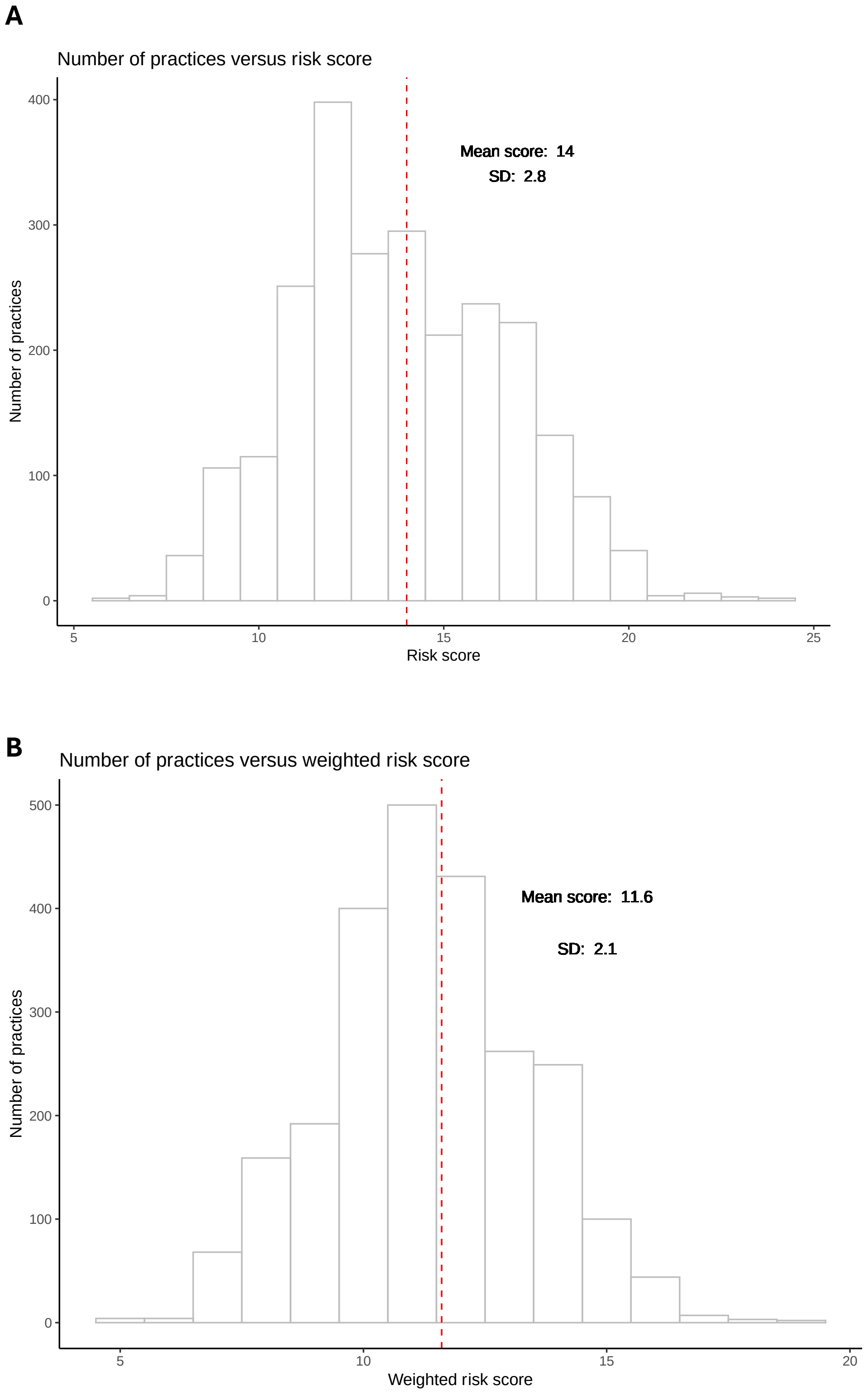


**Figure S3. Variations in risk scores distributions depending on categorical component weighting.** A: unweighted risk score distribution, all demographics. B: weighted risk score distribution, all demographics. Weighting was done as follows: 1 for all categorical components except for *Level of gregariousness* and *Immunocompetence*, which were given 0.5 because of their potentially lower impact on zoonotic pathogen spillover. Differences between weighted and unweighted scores were not significant.

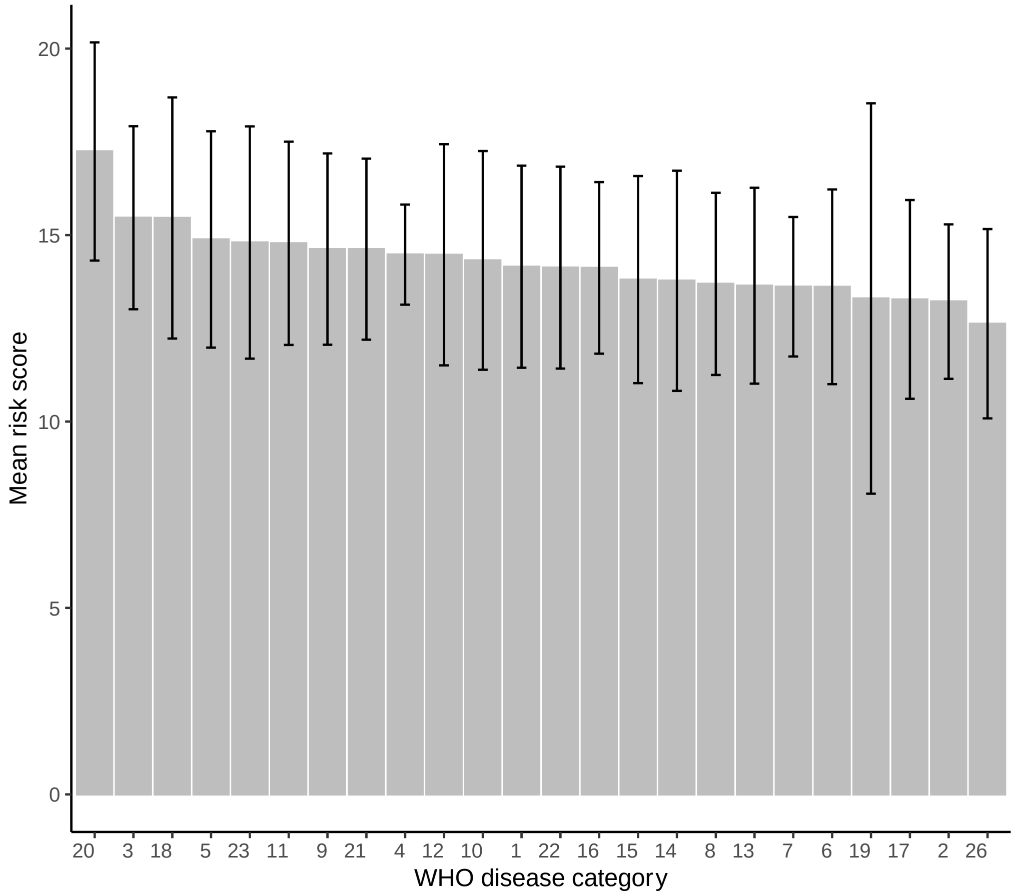


**Figure S4. Distribution of the risk score depending on the type of ailment treated.** (WHO disease categories displayed in Table S5)

**Table S5. WHO disease categories.** International Classification of Diseases by the World Health Organization (ICD-11 version; OMS 2019). Codes as in Table 1 in the main text.

| Codes | Diseases |
|---|---|
| 1 | Certain infectious or parasitic diseases |
| 2 | Neoplasms |
| 3 | Diseases of the blood or blood-forming organs |
| 4 | Diseases of the immune system |
| 5 | Endocrine, nutritional or metabolic diseases |
| 6 | Mental, behavioural or neuro-developmental disorders |
| 7 | Sleep-wake disorders |
| 8 | Diseases of the nervous system |
| 9 | Diseases of the visual system |
| 10 | Diseases of the ear or mastoid process |
| 11 | Diseases of the circulatory system |
| 12 | Diseases of the respiratory system |
| 13 | Diseases of the digestive system |
| 14 | Diseases of the skin |
| 15 | Diseases of the musculoskeletal system or connective tissue |
| 16 | Diseases of the genitourinary system |
| 17 | Conditions related to sexual health |
| 18 | Pregnancy, childbirth or the puerperium |
| 19 | Certain conditions originating in the perinatal period |
| 20 | Developmental anomalies |
| 21 | Symptoms, signs or clinical findings, not elsewhere classified |
| 22 | Injury, poisoning or certain other consequences of external causes |
| 23 | External causes of morbidity or mortality |
| 24 | Factors influencing health status or contact with health services |
| 25 | Codes for special purposes |
| 26 | Traditional Medicine conditions |
